# Elevated levels of hoarding in ADHD: a special link with inattention

**DOI:** 10.1101/2020.06.09.20126490

**Authors:** Sharon Morein-Zamir, Michael Kasese, Samuel R Chamberlain, Estherina Trachtenberg

**Author notes:** Sharon Morein, School of Psychology and Sport Science, Anglia Ruskin University, East Road, Cambridge CB1 1PT. Funding: The study was funded by the British Academy/Leverhulme (SG152110; PI:SMZ). SRC was funded by a Wellcome Trust Clinical Fellowship (reference 110049/Z/15/A).

## Abstract

Hoarding Disorder (HD) is under recognised and under-treated. Though HD develops by early adulthood, patients present only later in life, resulting in research based largely on samples of predominantly older females. Whilst formerly associated with Obsessive-Compulsive Disorder (OCD), it is now recognised that individuals with HD often have inattention symptoms reminiscent of Attention Deficit/Hyperactivity Disorder (ADHD). Here, we investigated HD in adults with ADHD. Patients in an ADHD clinic (n=88) reported on ADHD, HD and OCD-related symptoms, and compared with age, gender and education matched controls (n=90). Findings were assessed independently in an online UK sample to verify replication using a dimensional approach (n=220). Clinically significant hoarding symptoms were found in ~20% versus 2% of ADHD and control groups, respectively, with those with hoarding being on average in their thirties and with approximately half being male. Greater hoarding severity was noted even in the remaining patients compared with controls (d=0.89). Inattention was the only significant statistical predictor of hoarding severity in patients. Similarly, inattention, alongside depression and anxiety were the greatest predictors of hoarding in the independent sample where 3.2% identified as having clinically significant hoarding. Patients with ADHD had a high frequency of hoarding symptoms, which were specifically linked to inattention. HD should be routinely assessed in individuals with ADHD, as they do not typically disclose associated difficulties, despite these potentially leading to impaired everyday functioning. Research in HD should also investigate adults with ADHD, who are younger and with a greater prevalence of males than typical HD samples.

## Introduction

Individuals with Hoarding disorder (HD) suffer from excessive clutter, difficulties discarding and often excessive accumulation, causing clinically significant distress or impairment in social and occupational functioning (Frost and Hartl, 1996). Historically, hoarding symptoms were believed to characterise some Obsessive Compulsive Disorder (OCD) patients (Saxena, 2008). However, the majority of individuals with HD do not demonstrate the key features of OCD: obsessions or time-consuming compulsions (Frost et al., 2011). Consequently, since 2013 HD has been considered a distinct condition with unique phenomenological, psychological and neurobiological characteristics (American Psychiatric Association, 2013). HD is chronic with symptoms typically arising by late adolescence and steadily worsening over time, leading to significant distress and impact on daily functioning by age 40 (Grisham et al., 2006; Zaboski et al., 2019). With a prevalence of 2.5%, HD has similar rates for males and females (Postlethwaite et al., 2019). Notably, individuals with HD frequently have poor insight and consequently do not seek help or treatment (Frost et al., 2010; Grisham et al., 2005).

The bulk of hoarding research has drawn on patients with OCD, given its status prior to DSM-5. More recently, knowledge of HD *per se* and its treatment is derived from individuals who self-identify. These samples are predominantly female (Frost et al., 2011; Woody et al., 2020), suggesting insight may be especially poor in males. Moreover, HD participants in research and treatment studies are on average in their mid-50’s or 60’s (Tolin et al., 2015; Woody et al., 2014). Where community studies identify participants via housing, fire and public health agencies, gender is balanced but average age is even older (Woody et al., 2020). Taken together, this suggests that individuals, and particularly males who suffer from clinically significant hoarding already by their mid-30’s do not seek help until decades later, if at all.

Despite the historic association between HD and OCD as reflected in their nosology, research and clinical impression also point to a link between HD and Attention Deficit/Hyperactivity Disorder (ADHD). Patients with ADHD have persistent problems due to inattention and impulsivity, with over half experiencing functional impairments into adulthood (American Psychiatric Association, 2013). Notably, those with HD also have marked difficulties with attention, information processing and executive functioning (Morein-Zamir et al., 2014; Woody et al., 2014). This link was noted in individuals with OCD, with those with HD more likely to have ADHD, particularly the inattentive subtype (Frost et al., 2011; Sheppard et al., 2010). The association was also found in HD, with greater inattentive symptoms and elevated ADHD comorbidity rates compared to controls or patients with OCD (Frost et al., 2011; Hall et al., 2013; Hartl et al., 2005; Tolin and Villavicencio, 2011). A link between hoarding and impulsivity/hyperactivity has also been proposed, with HD patients endorsing more acquisition-related impulse-control problems (Frost et al., 2011). However, this association has proven inconsistent with positive findings in undergraduate and some patient samples (Hall et al., 2013; Hartl et al., 2005; Timpano et al., 2013), but not others (Sheppard et al., 2010; Tolin and Villavicencio, 2011). The link between hoarding and ADHD more generally was also not supported in a twin registry or an undergraduate sample (Ivanov et al., 2013; Woerner et al., 2017), though these studies assessed inattention and impulsivity/hyperactivity jointly.

To fully understand the association between hoarding and ADHD it is necessary to investigate hoarding in ADHD populations, with ADHD found in 2.5-3.5% of adults (Matte et al., 2015; Simon et al., 2009). A study in youths with ADHD highlighted high self-reported parental endorsement of clinically significant hoarding (29%), with inattention and hyperactivity/impulsivity independently predicting hoarding features (Hacker et al., 2016). An epidemiological study in adults using retrospective self-report also found a link between greater hoarding symptoms and childhood ADHD inattention, but not hyperactivity (Fullana et al., 2013).

Consideration of the link between hoarding and ADHD exists amongst hoarding specialists, but not amongst adult ADHD clinical and scientific specialists (Faraone et al., 2015; Posner et al., 2020). An investigation of hoarding in adult ADHD is needed to determine whether the link is present in this population which has been overlooked to date. Moreover, uncovering hoarding across early to middle-adulthood is key as this is when hoarding-related difficulties gradually amass to impairing levels. Discovery of elevated hoarding behaviors in this cohort may also promote earlier identification and intervention. This study addressed the gap by investigating patients at an adult ADHD clinic. In this well-characterised ADHD sample, we assessed frequency, severity and nature of hoarding symptoms in addition to OC-related traits and mood and anxiety (Hall et al., 2013). OC-related traits were assessed given their established link with hoarding. Perfectionism was also assessed as it has been previously associated with both OCD and hoarding (Pinto et al., 2017) but not ADHD. We also explored differences in clinical characteristics between those with clinically significant hoarding relative to non-hoarding patients. In view of the inconsistencies in the literature, we examined whether inattention and impulsivity/hyperactivity uniquely contributed to hoarding. We compared the adult ADHD sample with a control group matched for age, gender and education. An independent community sample allowed us to verify whether key findings replicated using a similar procedure when a dimensional approach to hoarding and ADHD behaviors was adopted (Morein-Zamir et al., 2020).

## Materials and Methods

### Participants

Patients with ADHD were recruited from the NHS Cambridge and Peterborough Foundation Trust Adult ADHD clinic. Diagnoses in clinic were according to DSM-5 (American Psychiatric Association, 2013) and based on a full structured clinical interview with the patient and an informant who had known them since childhood. Assessments also identified comorbidities and were conducted by a clinical professional with extensive expertise in ADHD assessment. Clinical assessment also included Barkley Adult ADHD Rating Scales, self- and informant-report of childhood and adulthood symptoms (BAARS; (Barkley, 2011)).

Control participants were recruited via advertisements in the local Cambridge community. Screening verified controls did not meet diagnostic threshold for ADHD and included a validated brief ADHD screener (Kessler et al., 2005; Ustun et al., 2017) in addition to the MINI (Sheehan et al., 1998). Exclusion criteria for controls included current or former ADHD diagnosis, or probable ADHD on the screener, with screening verifying ongoing appropriate sampling of age, gender and education levels to match the patients. Exclusion criteria for all participants above included severe neurological deficits. Patients were contacted by mail and in clinic to participate in a study about “Accumulation behaviors in ADHD”, with a 32% response rate. Principle recruitment and testing occurred from July 2017 to August 2018. For the independent UK online cohort (recruited via Prolific Academic, prolific.co.uk in November 2018) there were no exclusion criteria, with the provision that the sample be 50% female.

### Measures

#### ADHD and impulsivity

The Adult ADHD Self-Report Scale (ASRS; (Kessler et al., 2005)) assesses severity of the 18 DSM-IV symptoms of inattention, hyperactivity and impulsivity. ASRS scores have shown good reliability and validity in clinical and general populations (Brevik et al., 2020). Subscale and total scores in current samples demonstrated high internal consistency (Cronbach α values 0.81-0.95). For patients, the BAARS-IV self-report was available, which includes 27 items describing behavior over the past 6 months, with subscales for inattention, hyperactivity, impulsivity and sluggish cognitive tempo. The scale had good internal consistencies in the present sample (subscale α values 0.80-0.92). The widely used Barret Impulsivity Scale-11 (BIS; (Patton et al., 1995)) comprises 30 items describing impulsive or non-impulsive (reverse scored) behaviors and preferences. Given its inconsistent factor structure (Reise et al., 2013), only total score was calculated with good current internal consistency (α values 0.92 and 0.83).

#### Hoarding and clutter

The Savings Inventory Revised (SIR, (Frost et al., 2004)) contains 23 items and three subscales: difficulty discarding, excessive acquisition and clutter, with a clinical cut-off of 41 (Frost et al., 2011). The SIR has demonstrated excellent internal consistency, good test-retest reliability and convergent validity with current samples showing excellent internal consistency on all subscales (α values 0.84-0.96). The Clutter Image Rating Scale (CIR; (Frost et al., 2008)) presents nine photographs for each room (living room, kitchen, and bedroom) with increasing clutter and participants choose the most closely representing their own. Another room was shown (bathroom) but not analysed further. CIR has demonstrated good psychometric properties with current samples having good internal consistency (current α values 0.76 and 0.83)

#### Obsessive Compulsive severity

The Obsessive Compulsive Inventory-Revised (OCIR; (Foa et al., 2002)) is widely used to assess OCD traits with 18 items on hoarding, checking, washing, ordering, obsessing, and neutralising subscales. The OCIR has good internal consistency, test-retest reliability and convergent and discriminant validity (Foa et al., 2002), with current subscales showing good to excellent consistency in both samples (α values 0.78-0.94).

#### Perfectionism

The Frost Multidimensional Perfectionism Scale (MPS; (Frost et al., 1990)) contains 35 items generating six subscales: concern over mistakes, doubting of actions, personal standards, parental expectations, parental criticism, and organization. It is reliable and valid for non-clinical and clinical populations (Frost et al., 1990). Current subscales showed good to excellent internal consistency in both samples (α values 0.78-0.92).

#### Depression and anxiety

Depression and Anxiety Severity Scale (DASS; (Lovibond and Lovibond, 1995)) contains 21 items with participants rating items over the past week. The DASS has demonstrated excellent internal consistency and concurrent validity, as did total scores for present samples (α=0.95 for both).

#### Functional impairment

The Weiss functional Impairment Scale (WFIRS; (Weiss, 2005)) contains 70 items with participants rating the extent to which emotional and behavioral problems have affected 7 domains over the last month (family, work, school, life skills, self-concept, social and risk). Higher mean scores indicate greater functional impairment. Previous studies have shown good psychometric characteristics (Canu et al., 2016), and current samples had good internal consistency in all domains (α values 0.74-0.93).

### Procedure

Eligible and interested participants were sent a link to an online survey, with the data for patients who consented supplemented with in-clinic data (BAARS, WFIRS). The survey consisted of a demographics section followed by questionnaires presented in random order (ASRS, SIR, CIR, BIS, DASS, OCIR, and MPS). All participants provided informed consent before taking part in accordance with the Declaration of Helsinki and were compensated. The study was approved by an NHS Research Ethics Committee (16/WM/0368) and received Health Research Authority approval. The subsequent online study was approved by the University Ethics Panel. Testing procedures for the online sample were identical, with the following exceptions. Four attention check items were included to identify careless responding (Meade and Craig, 2012). Additional questionnaires were administered but not reported here.

### Analyses and Design

Group comparisons used Chi-squared and Mann-Whitney tests for categorical and continuous variables, respectively. Based on previous literature (Frost et al., 2008; Tolin et al., 2010; Wootton et al., 2015), clinically meaningful hoarding symptoms was defined as SIR values greater than 40 *and* OCIR-Hoarding greater than 6 *and* a rating of 4 in at least one CIR room. CIR average of 4 (Frost et al., 2008) would have unduly focused on clutter and appeared overly stringent given diagnosed HD samples have lower values (Tolin et al., 2010). Participant frequency above and below the threshold was compared between the ADHD and control samples. Further comparisons assessed group differences in those below the threshold, and within the ADHD group between those above versus below the threshold.

Clinic-based data for the BAARS self-report was available for 59 patients and WFIRS for 58. Where analyses do not include the full sample, the number of observations is reported. Where multiple tests were performed, Bonferroni correction for type-I error was applied. Finally, we examined the contribution of ADHD, depression and anxiety and OC-related variables to the presence of hoarding symptoms. This was done separately in each group given multi-collinearity with ADHD symptoms and group differences in SIR, and then verified separately in the online sample. OCIR subscales were included as we hypothesised they constitute distinct behavioral dimensions in ADHD. Regression models using bootstrapped 95% confidence intervals (CI) and model parameters are reported, with variance inflation factor (VIF) values used to inspect multicolinearity and Cook’s Distance to inspect the influence of individual data points. Values were acceptable, with VIF scores of less than 2.1 for all models.

## Results

The ADHD and control groups did not differ significantly in gender, age and family status but did in living arrangements (see Table 1). As expected given their clinical diagnosis, patients scored significantly higher on all ADHD symptom severity measures, in addition to higher levels of depression and anxiety. In-clinic data indicated that the most common comorbidities in the ADHD group were depression and anxiety (18%) followed by Autism Spectrum Disorder (11.3%), OCD (4.5%), and eating disorders including binge eating disorder (3.4%). One patient was diagnosed with HD. Compared with controls, patients reported greater levels in all hoarding related questionnaires and subscales, all of which survived Bonferroni correction (see also Figure 1). Several additional OCIR-subscales were higher in patients with ADHD, though only OCIR-obsessing survived multiple comparisons correction. Several perfectionism subscales pointed to group differences, with patients reporting lower organisation and greater concern over mistakes, doubting of actions and parental criticism, though the later did not survive correction.

**Table 1.** Demographic and clinical characteristics of ADHD patient and control groups

| Characteristic | ADHD<br>(n=88) | Controls<br>(n=90) | $Z/\chi^2$ | $d$ | $p$ |
| --- | --- | --- | --- | --- | --- |
|  | Mean (SD) | Mean (SD) |  |  |  |
| Age in years | 32.68 (10.31) | 31.99 (10.82) | 0.56 |  | .55 |
| Gender | 56:32 | 58:32 | 0.01 |  | .91 |
| M:F |  |  |  |  |  |
| Education |  |  | 5.14 |  | .162 |
| GCSE/O levels | 24 | 14 |  |  |  |
| A-levels/Equivalent | 11 | 12 |  |  |  |
| Undergraduate | 31 | 34 |  |  |  |
| Post graduate | 21 | 30 |  |  |  |
| Living with: |  |  | 10.33 |  | .006 |
| Alone | 13 | 13 |  |  |  |
| Non-family | 5 | 20 |  |  |  |
| Family | 69 | 56 |  |  |  |
| Family Status |  |  | 1.98 |  | .74 |
| Single | 33 | 31 |  |  |  |
| Separated/divorced | 4 | 3 |  |  |  |
| Partnered -living alone | 7 | 11 |  |  |  |
| Living with partner<br>/married | 44 | 45 |  |  |  |
| Members in Household | 3.11 (1.63) | 3.16 (1.61) | 0.27 |  | .78 |
| Inattention (ASRS) | 28.92 (5.21) | 14.06 (5.94) | 10.53 | 2.66 | <.001 |
| Hyperactivity/<br>impulsivity (ASRS) | 24.14 (6.38) | 11.89 (6.19) | 9.45 | 1.95 | <.001 |
| Impulsivity (BIS) | 83.06 (11.29) | 60.50 (10.61) | 9.76 | 2.06 | <.001 |
| Clutter (SIR) | 13.48 (7.19) | 7.56 (4.65) | 5.72 | .98 | <.001 |
| Difficulty Discarding<br>(SIR) | 9.88 (6.06) | 4.86 (3.95) | 5.79 | .98 | <.001 |
| Acquisition (SIR) | 10.24 (5.91) | 4.92 (4.14) | 6.09 | 1.04 | <.001 |
| Hoarding (SIR Total) | 33.59 (18.15) | 17.33 (12.09) | 6.15 | 1.06 | <.001 |
| Clutter (CIR) | 2.26 (1.01) | 1.61 (0.59) | 4.34 | .68 | <.001 |
| Hoarding (OCIR) | 4.94 (3.43) | 2.81 (2.72) | 4.25 | .69 | <.001 |
| Washing (OCIR) | 2.12 (3.05) | 1.52 (2.34) | 0.53 | 0.22 | .56 |
| Neutralizing (OCIR) | 2.15 (2.92) | 1.56 (2.43) | 0.61 | 0.22 | .52 |
| Checking (OCIR) | 3.76 (3.38) | 2.39 (2.56) | 2.61 | 0.46 | .008 |
| Ordering (OCIR) | 4.48 (3.69) | 3.14 (3.11) | 2.37 | 0.39 | .017 |
| Obsessing (OCIR) | 5.46 (3.78) | 1.88 (2.37) | 6.66 | 1.14 | <.001 |
| OCIR-Total | 22.92 (14.73) | 13.30 (11.86) | 4.62 | 0.68 | <.001 |
| Concern Over Mistakes<br>(MPS) | 28.95 (9.15) | 22.32 (7.22) | 4.85 | 0.81 | <.001 |
| Personal Standards (MPS) | 22.83 (6.67) | 22.39 (4.98) | 0.81 | 0.08 | 0.42 |
| Parental Expectations (MPS) | 13.97 (4.77) | 12.93 (4.59) | 1.32 | 0.22 | 0.19 |
| Parental Criticism (MPS) | 10.81 (4.84) | 8.84 (3.32) | 2.51 | 0.47 | 0.012 |
| Doubting of Actions (MPS) | 14.99 (3.39) | 11.23 (2.95) | 6.89 | 1.18 | <.001 |
| Organisation (MPS) | 18.08 (5.84) | 21.43 (5.04) | 3.83 | 0.62 | <.001 |
| Depression & Anxiety (DASS) | 27.51 (13.55) | 10.30 (8.65) | 8.27 | 1.52 | <.001 |

**Note.** ASRS= Adult ADHD Self-Report Scale; BIS=Barratt Impulsivity Scale; CIR=Clutter Image Rating Scale; OCIR=Obsessive Compulsive Inventory Revised; MPS-Frost Multidimensional Perfectionism Scale; DASS=Depression and Anxiety Scale.

**Figure 1.**
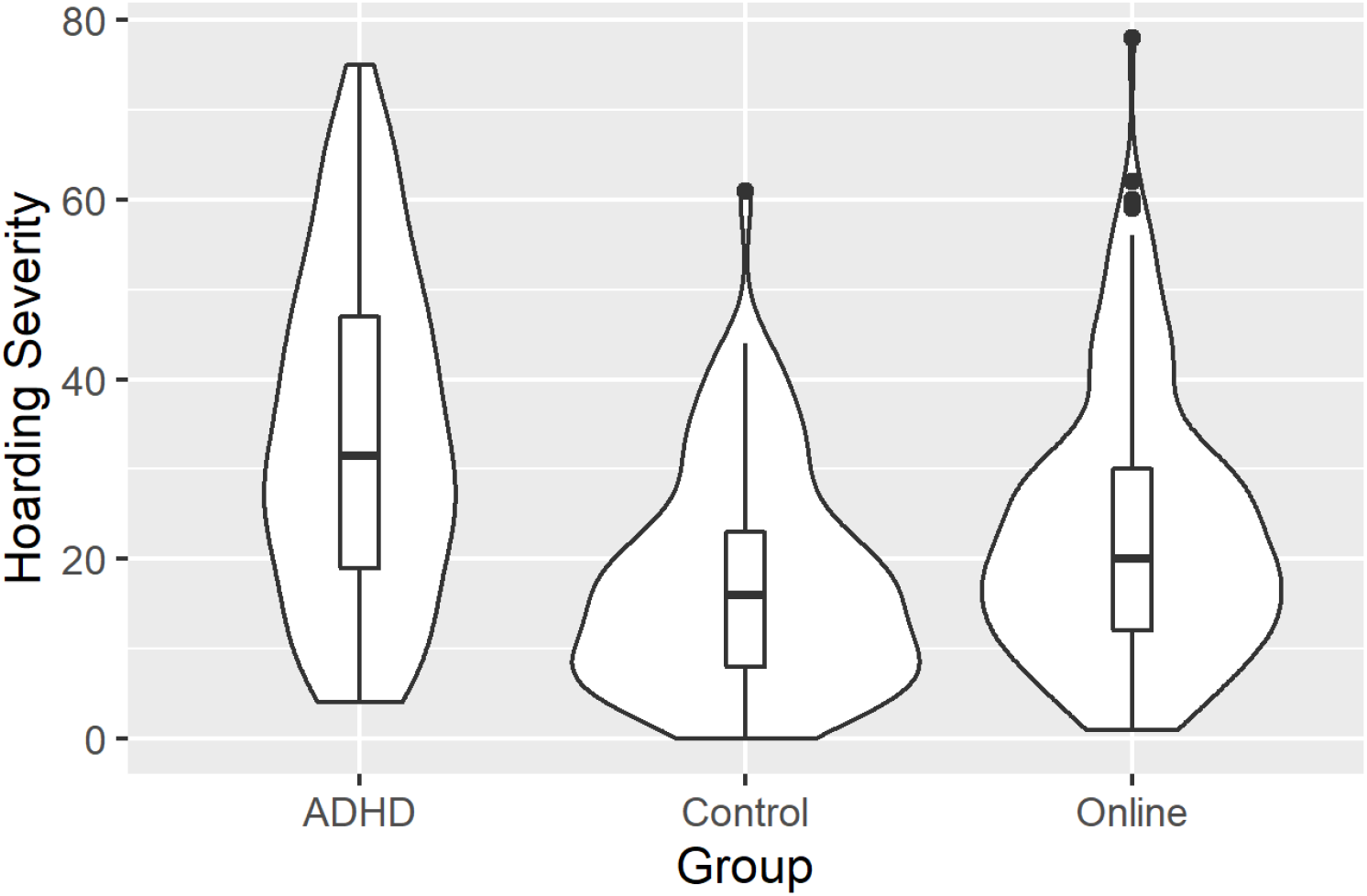
Violin plots showing hoarding severity (Savings Inventory Revised total) for the ADHD, control and online samples. The box-plots denote median and inter-quartile range and are overlaid with a density curve conveying the distribution shape of the data.

There were 17 patients and 2 controls who scored above the threshold of clinically significant hoarding (19.31% vs. 2.22%, χ^2^(1)=13.64, p<.001). Elevated hoarding in the SIR was also found in patients who were below the threshold compared to controls, (27.69, n=71 vs. 16.57, n=88, Z=4.91, p<.001, d=0.88). Similarly, these patients had higher OCI-hoarding compared with controls (3.87 vs. 2.66, Z=2.81, p=.004, d=0.45) and higher CIR values (1.92 vs. 1.57, Z=2.34, p=.004, d=0.55).

Within the ADHD group, we inspected whether patients with clinically significant hoarding symptoms differed from the remaining patients. We did not observe a significant age difference with all being in their 30’s on average (36.17 vs. 31.84, Z=1.51, p=.132). There was no significant gender difference (χ^2^(1)=0.01, p=.92), with 6 females of the 17 above threshold. There was also no difference in overall medication status with 71% and 82% of those above and below threshold being medicated respectively (χ^2^(1)=1.04, p=.31). Nor did there seem to be significant patterns in medication type (e.g., atomoxetine specifically) though a greater proportion above the cut-off were receiving SSRIs (47% vs 27%, p=.10). Patients above the threshold reported significantly greater levels of ASRS-inattention (32.41 vs. 28.08, Z=3.54, p<.001, d=0.88) but not ASRS-hyperactivity/impulsivity or BIS impulsivity. This was consistent with their worse inattention symptoms in the BAARS-inattention subscale (31.29, n=14, vs. 24.98, n=45, Z=2.90, p=.003, d=0.90). Whilst those above the cut-off also had significantly greater BAARS-hyperactivity (Z=2.05, p=.04), this did not survive correction nor were there differences in BAARS-impulsivity or cognitive tempo. Additionally, those above threshold reported greater depression and anxiety (37.71 vs. 25.07, Z=3.00, p=.002, d=1.00), as well as concerns over mistakes (34.29 vs. 27.67, Z=2.85, p=.003) and parental criticism (13.71 vs. 10.11, Z=2.77, p=.005). WFIRS mean functional impairment was higher for the patients above compared to below the threshold (1.98, n=13 vs. 1.19, n=47, Z=3.90, p<.001).

### Associations with hoarding severity

Associations between ADHD symptoms and hoarding indices was examined in the ADHD sample (Table 2). ASRS-inattention was consistently positively associated with all five hoarding indices whilst ASRS-hyperactivity/impulsivity was only associated with clutter. Secondary analyses on the BAARS revealed a similar pattern. BAARS-inattention and BAARS-cognition were significantly associated with all three SIR subscales (r_s_ values of 0.29-0.30) whereas BAARS-hyperactivity was significantly associated with SIR-clutter only (r_s_=0.32) and BAARS-impulsivity was not significantly associated with any SIR subscale. In contrast, in controls both ASRS-inattention and ASRS-hyperactivity were positivity associated with most hoarding indices.

**Table 2.**
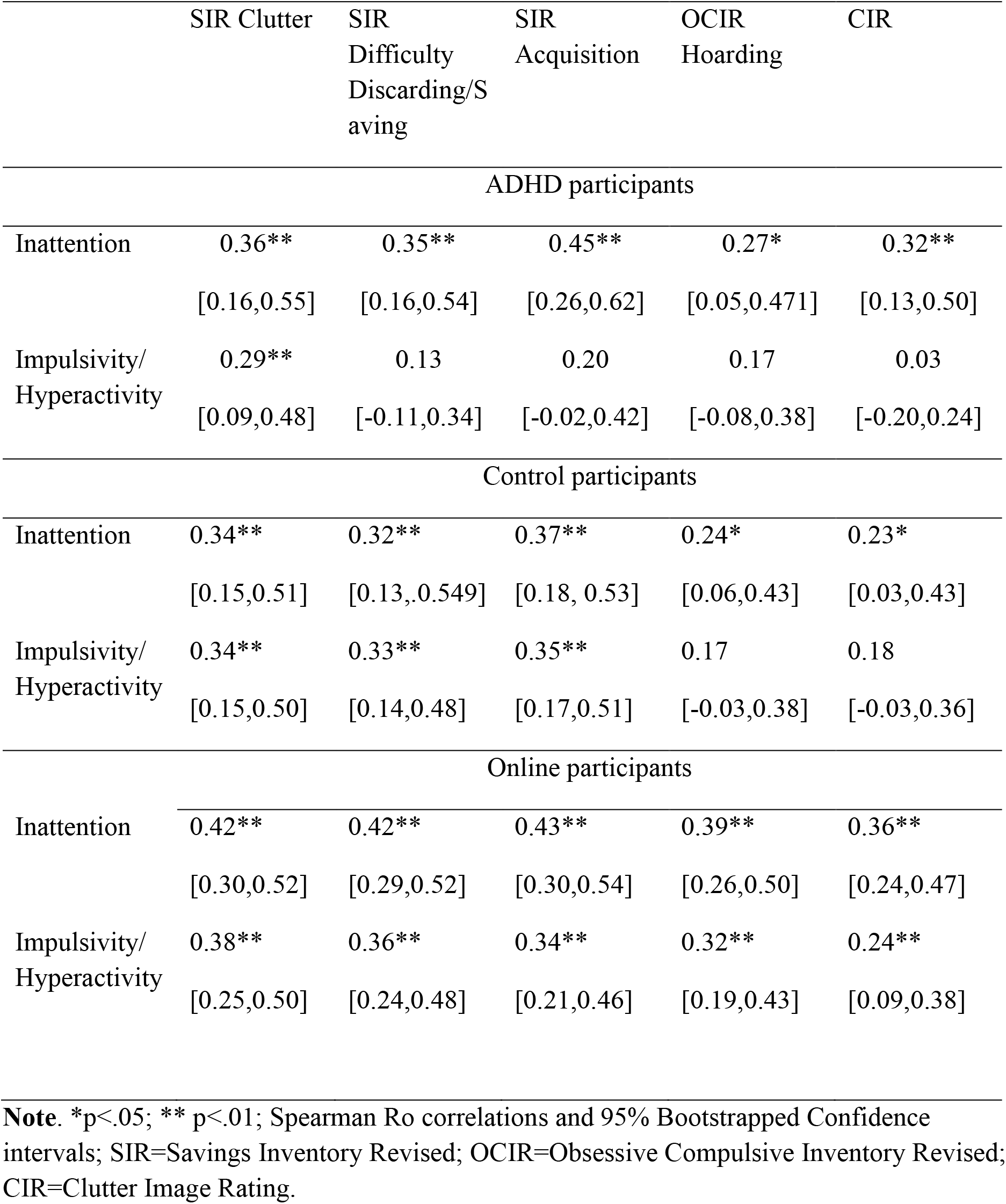
Spearman Ro’s correlations between ASRS subscales and hoarding-related indices in each of the three groups.

### Statistical predictors of hoarding severity

In patients, a regression analysis with hoarding (SIR total) as outcome and ADHD symptom subscales, BIS-impulsivity, anxiety and depression, and OC-related symptoms as predictors (R^2^=0.33) found only inattention to be a significant predictor (Table 3). In controls this analysis revealed checking and to a lesser degree neutralizing as significant predictors (R^2^=0.37). Similar regression models including gender and age in both groups or without BIS-impulsivity given concerns over multicollinearity yielded the same results. Additional models indicated similar conclusions for each hoarding subscale when considered separately.

**Table 3.**
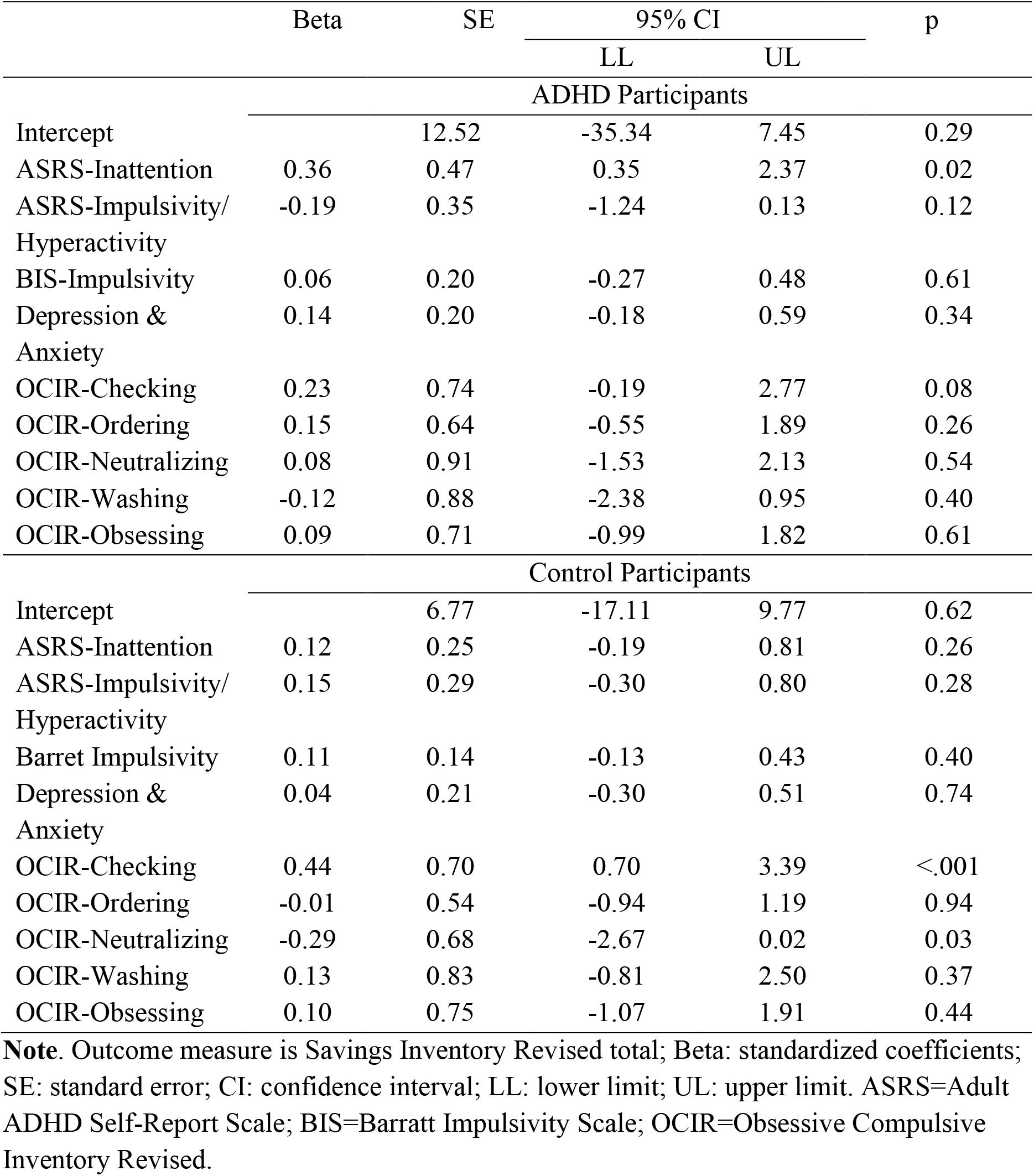
Results of regression analyses for ADHD and control participants

|  | Beta | SE | 95% CI |  | p |
| --- | --- | --- | --- | --- | --- |
|  |  |  | LL | UL |  |
| ADHD Participants |  |  |  |  |  |
| Intercept |  | 12.52 | -35.34 | 7.45 | 0.29 |
| ASRS-Inattention | 0.36 | 0.47 | 0.35 | 2.37 | 0.02 |
| ASRS-Impulsivity/<br>Hyperactivity | -0.19 | 0.35 | -1.24 | 0.13 | 0.12 |
| BIS-Impulsivity | 0.06 | 0.20 | -0.27 | 0.48 | 0.61 |
| Depression &<br>Anxiety | 0.14 | 0.20 | -0.18 | 0.59 | 0.34 |
| OCIR-Checking | 0.23 | 0.74 | -0.19 | 2.77 | 0.08 |
| OCIR-Ordering | 0.15 | 0.64 | -0.55 | 1.89 | 0.26 |
| OCIR-Neutralizing | 0.08 | 0.91 | -1.53 | 2.13 | 0.54 |
| OCIR-Washing | -0.12 | 0.88 | -2.38 | 0.95 | 0.40 |
| OCIR-Obsessing | 0.09 | 0.71 | -0.99 | 1.82 | 0.61 |
| Control Participants |  |  |  |  |  |
| Intercept |  | 6.77 | -17.11 | 9.77 | 0.62 |
| ASRS-Inattention | 0.12 | 0.25 | -0.19 | 0.81 | 0.26 |
| ASRS-Impulsivity/<br>Hyperactivity | 0.15 | 0.29 | -0.30 | 0.80 | 0.28 |
| Barret Impulsivity | 0.11 | 0.14 | -0.13 | 0.43 | 0.40 |
| Depression &<br>Anxiety | 0.04 | 0.21 | -0.30 | 0.51 | 0.74 |
| OCIR-Checking | 0.44 | 0.70 | 0.70 | 3.39 | <.001 |
| OCIR-Ordering | -0.01 | 0.54 | -0.94 | 1.19 | 0.94 |
| OCIR-Neutralizing | -0.29 | 0.68 | -2.67 | 0.02 | 0.03 |
| OCIR-Washing | 0.13 | 0.83 | -0.81 | 2.50 | 0.37 |
| OCIR-Obsessing | 0.10 | 0.75 | -1.07 | 1.91 | 0.44 |

**Note.** Outcome measure is Savings Inventory Revised total; Beta: standardized coefficients; SE: standard error; CI: confidence interval; LL: lower limit; UL: upper limit. ASRS=Adult ADHD Self-Report Scale; BIS=Barratt Impulsivity Scale; OCIR=Obsessive Compulsive Inventory Revised.

### Independent online sample findings

The sample consisted of 220 individuals (111 female), with a mean age of 35.78 (SD=12.74). An additional four were removed having failed the attention checks. Clinically significant hoarding symptoms were found in 8 individuals (3.64%). Mean SIR clutter, discarding and acquisition values were 9.68 (SD=5.36), 6.65 (SD=4.64) and 6.61 (SD=4.81), respectively. Mean OCI-hoarding was 3.28 (SD=2.86) and mean CIR was 1.78 (SD=0.73). The association between ADHD and hoarding indices pointed to consistent medium to large associations with hoarding for both inattention and for hyperactivity/impulsivity (see Table 2). A regression analysis with SIR total as outcome and ADHD symptoms, BIS-impulsivity, anxiety and depression, and OC-related subscales as predictors found a good fit (R^2^=0.36). Again inattention was a significant predictor, in addition to depression and anxiety and ordering (Table 4).

**Table 4.**
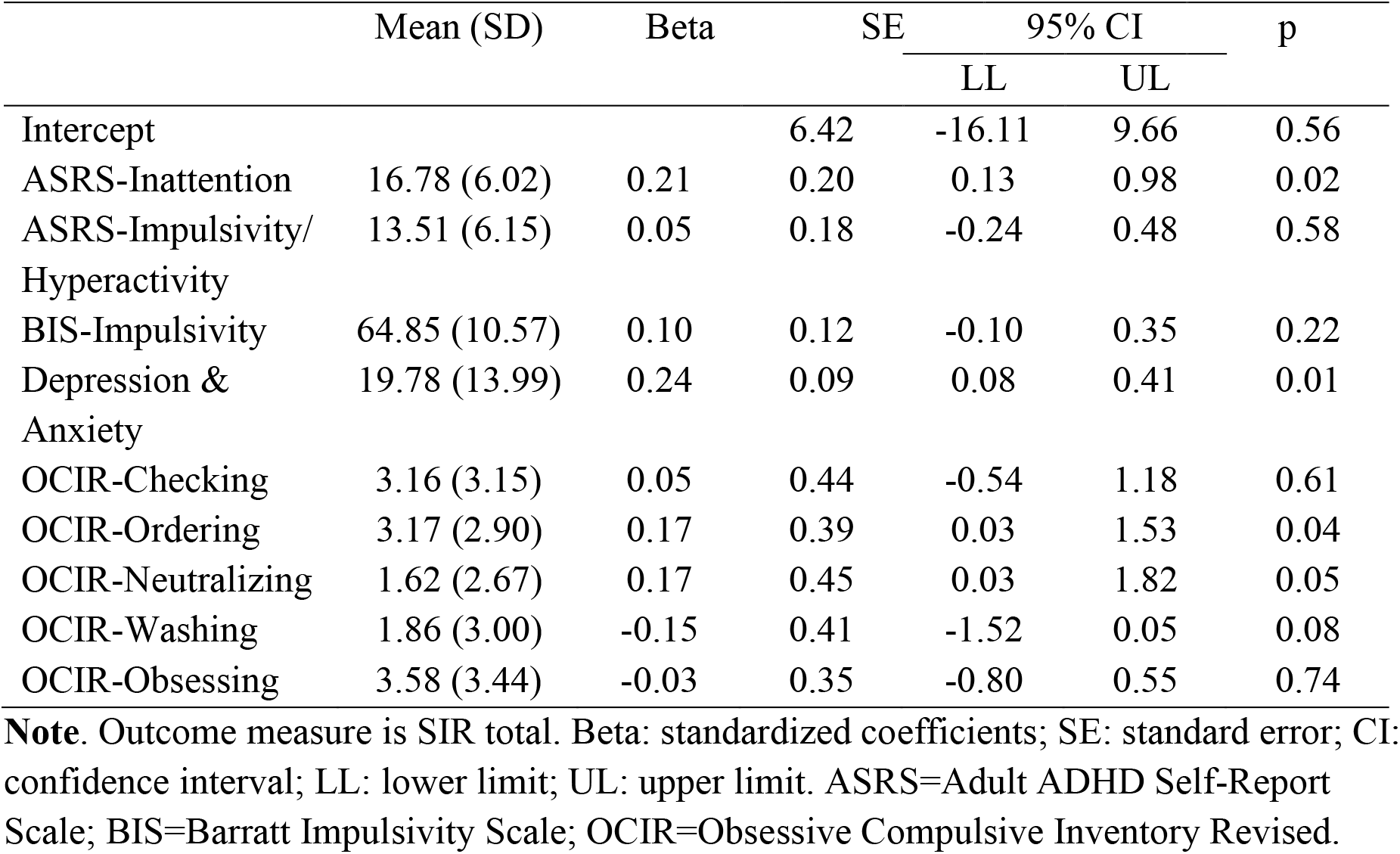
Online UK sample descriptive statistics and results of regression analysis.

|  | Mean (SD) | Beta | SE | 95% CI |  | p |
| --- | --- | --- | --- | --- | --- | --- |
|  |  |  |  | LL | UL |  |
| Intercept |  |  | 6.42 | -16.11 | 9.66 | 0.56 |
| ASRS-Inattention | 16.78 (6.02) | 0.21 | 0.20 | 0.13 | 0.98 | 0.02 |
| ASRS-Impulsivity/<br>Hyperactivity | 13.51 (6.15) | 0.05 | 0.18 | -0.24 | 0.48 | 0.58 |
| BIS-Impulsivity | 64.85 (10.57) | 0.10 | 0.12 | -0.10 | 0.35 | 0.22 |
| Depression &<br>Anxiety | 19.78 (13.99) | 0.24 | 0.09 | 0.08 | 0.41 | 0.01 |
| OCIR-Checking | 3.16 (3.15) | 0.05 | 0.44 | -0.54 | 1.18 | 0.61 |
| OCIR-Ordering | 3.17 (2.90) | 0.17 | 0.39 | 0.03 | 1.53 | 0.04 |
| OCIR-Neutralizing | 1.62 (2.67) | 0.17 | 0.45 | 0.03 | 1.82 | 0.05 |
| OCIR-Washing | 1.86 (3.00) | -0.15 | 0.41 | -1.52 | 0.05 | 0.08 |
| OCIR-Obsessing | 3.58 (3.44) | -0.03 | 0.35 | -0.80 | 0.55 | 0.74 |

**Note.** Outcome measure is SIR total. Beta: standardized coefficients; SE: standard error; CI: confidence interval; LL: lower limit; UL: upper limit. ASRS=Adult ADHD Self-Report Scale; BIS=Barratt Impulsivity Scale; OCIR=Obsessive Compulsive Inventory Revised.

## Discussion

The findings show significantly elevated hoarding symptoms in adult individuals with ADHD compared to controls matched for key demographic characteristics. Clinically significant levels of hoarding were reported by approximately 20% of ADHD patients, similar to levels previously reported in OCD cohorts (Frost et al., 2011). Patients who did not have clinically significant hoarding still reported more symptoms than controls, with a large effect size. Present clinical and anecdotal evidence indicated limited insight and that patients did not spontaneously raise hoarding-related issues, though they did endorse them once explicitly questioned. The results suggest that hoarding symptoms should be routinely assessed in patients with ADHD, particularly given limited awareness of impairments associated with them. Clinically significant hoarding was associated with worse quality of life and higher depression and anxiety. Depression in HD stems in part from the social, familial and occupational issues that emerge from chronic hoarding (Tolin et al., 2008). It may also be linked to emotional dysregulation which is routinely noted in both HD and in ADHD and itself contributes to impairment (Frost et al., 2011; Surman et al., 2013).

Consideration of comorbid hoarding in ADHD should contribute to behavioral and pharmacotherapy treatment choices. For example, preliminary open-label data suggest atomoxetine may be effective for HD symptoms (Grassi et al., 2016), as do some SSRIs (Piacentino et al., 2019). However, in part due to HD being recognised only recently randomized control trials are still lacking. In any case, greater awareness amongst clinicians, patients and their carers about the link between ADHD and hoarding could also facilitate more effective long-term management, as hoarding often gradually worsens with time. Further research into evidence-based treatment options for HD, including when co-presenting with ADHD or in mid-adulthood, is urgently needed.

Present findings point to a robust and unique association between hoarding and inattention extending previous OCD and HD studies to patients with ADHD (Hall et al., 2013; Hartl et al., 2005; Sheppard et al., 2010). Individuals with ADHD and clinically meaningful hoarding had more severe inattention symptoms compared to those below the cut-off with large effect sizes on two independent measures taken at different time points. The unique contribution of inattention to hoarding was replicated in an independent sample generalising the findings to subclinical populations and supporting the dimensionality of hoarding symptoms. Consistent associations between inattention and hoarding were also found in controls. The association is underscored by inattention and executive dysfunction contributing to symptoms in both disorders. Some patients with ADHD show functional deficits of impaired organization, planning and working memory (Faraone et al., 2015; Nigg, 2005). These difficulties may jointly underscore both ADHD and HD symptoms, suggesting that those with ADHD who experience executive dysfunction should suffer most from hoarding symptoms. This would anticipate that high heritability found in both (Faraone and Larsson, 2019; Iervolino et al., 2009) could stem in part from shared genetic vulnerability (Hirschtritt et al., 2018).

Despite some overlap, inattention and impulsivity/hyperactivity are separate domains with domain specific genetic influences (Faraone et al., 2015). A unique link between impulsivity/hyperactivity and hoarding was not found in any of the present cohorts suggesting the observed correlations were driven by general psychopathology-related variance. This is consistent with findings from OCD samples (Sheppard et al., 2010; Tolin and Villavicencio, 2011). The link should be assessed in different populations, for example it is still possible hoarding is associated with impulsivity specifically in HD, possibly expressed by impulse control problems (Hall et al., 2013; Hartl et al., 2005). The absence of the unique association contrasts with its presence in children with ADHD (Hacker et al., 2016) and may stem from development trajectories relating to impulsivity/hyperactivity and hoarding. We also noted that patients with ADHD scored higher than controls on some but not all OCIR and perfectionism subscales. ADHD appears to be associated with a unique profile, which could stem from ADHD-specific symptoms in addition to the well-established association between ADHD and OCD. Namely, some patients with ADHD may endorse checking, doubts about actions, concern over mistakes and parental criticism due to the lifelong consequences of having ADHD. Also in light of concerns over discriminative validity of items relating to OC and inattention (Morein-Zamir et al., 2020), interpreting total values on OCIR and perfectionism scales in this population should be treated with caution.

Current findings also highlight the presence of patients with hoarding who were on average in their thirties. There was no gender difference in the ADHD sample, nor was gender associated with hoarding severity in the other samples. Patients above the hoarding cut-off were closer in demographic profile to that expected in the population compared to typical research studies (Woody et al., 2020). This implies that HD research should actively seek adult ADHD participants with hoarding given their younger age and greater prevalence of males. It remains to be established whether the clinical and neuropsychological profiles of such individuals differ from typical HD cohorts who self-identify or those with comorbid OCD. Already some evidence points to cognitive and neural differences in HD depending on the presence of comorbid OCD (Hough et al., 2016; Mataix-Cols et al., 2004; Morein-Zamir et al., 2014). Taken together, more flexible theoretical HD models alongside more targeted behavioral treatments may be needed (Lynch et al., 2017).

The present study did not assess the phenomenology of hoarding, such as the presence of a strong emotional and sentimental attachment to possessions (Frost and Hartl, 1996; Grisham et al., 2009). This could be a key difference between those with hoarding with versus without ADHD, with symptoms in the former more closely linked to cognitive dysfunction (Fullana et al., 2013; Hacker et al., 2016). However qualitative data suggest some children with ADHD have a strong sentimental attachment with increased distress when confronted with the need to discard (Lynch et al., 2017). Along with the presence of emotional dysregulation as mentioned above, hoarding in ADHD does not appear to be simply a consequence of inattention or impulsivity (Lynch et al., 2015). Despite the overlap between ADHD and HD, most ADHD participants did not report clinically meaningful hoarding though many had subclinical levels. This highlights the dimensionality of hoarding symptoms and is consistent with contributing independent pathways. For example, hoarding appears elevated in individuals with autism, anorexia nervosa (Halmi et al., 2003; Storch et al., 2016) and anxiety disorders (Tolin et al., 2011). However, it is presently unclear whether increased hoarding in these disorders is underscored by concomitant raised levels of ADHD or OCD symptoms as found in Tourette’s syndrome (Huisman-van Dijk et al., 2016).

The present study relied on self-report rather than clinician-rated measures of hoarding with the study information mentioning ‘accumulation behaviors’, allowing potential biasing in the ADHD participants. However, the control group was provided with identical information and experienced the same procedures mitigating this concern, as did the replication in the online sample of the link with inattention. Additionally, multiple well-validated measures for ADHD and HD-related symptoms were collected, while employing previously established cut-off values for HD. This allowed us to avoid an overly liberal approach to hoarding difficulties. Moreover, present criteria yielded an online sample prevalence within the confidence intervals of a recent meta-analyses (Postlethwaite et al., 2019).

In sum, this study points to a hidden population of adults who demonstrate clinically significant hoarding symptoms. Inattention symptoms were specifically linked to hoarding severity and this was replicated in an independent online sample. The results suggest hoarding symptoms should be assessed in patients with ADHD, particularly given insight issues. Moreover, HD research should actively investigate hoarding in such individuals given their age and gender ratio. A better understanding of the overlap between HD and ADHD will enrich theorizing and treatment development to ultimately improve functioning and outcomes for all.

## Data Availability

Data are available from first author on request.

## Acknowledgements

We wish to thank all participants and the staff at the Adult ADHD Clinic in the Cambridge and Peterborough Foundation Trust, in addition to Vicki Slater and Natalia Rymarcyzk who assisted in data collection.

